# Plasma Proteomics and Sensitive Imaging Biomarkers of Vascular Brain Injury

**DOI:** 10.64898/2026.01.19.26344369

**Authors:** Qiong Yang, Valborg Gudmundsdottir, Emer R. McGrath, Muralidharan Sargurupremraj, Yineng Zhu, Pauline Maillard, Zsu-Zsu Chen, Claudia L. Satizabal, Usman Tahir, Alexa S Beiser, Joseph Loureiro, Lars Forsberg, Stephanie Debette, Charles DeCarli, Lenore Launer, Ramachandran S Vasan, Lori L. Jennings, Robert E Gerszten, Sudha Seshadri, Vilmundur Guðnason

**Author notes:** Corresponding Author: Qiong Yang, PhD, Department of Biostatistics, Boston University School of Public Health 801 Mass Ave, Boston, MA 02118. These authors contributed equally (Qiong Yang, Valborg Gudmundsdottir). These authors contributed equally (Sudha Seshadri, Vilmundur Guðnason).

## Abstract

**Background and Objectives:** Identifying proteomic biomarkers associated with neuroimaging endophenotypes of cerebrovascular dysfunction and generalized neurodegeneration may provide insights into pre-Alzheimer disease pathological processes. This study aims to examine a broad panel of circulating proteins in relation to two diffusion tensor imaging (DTI) measures:free water (FW) and peak width of skeletonized mean diffusivity (PSMD), which are sensitive indicators of early white matter injury.

**Methods:** Using data from the Framingham Heart Study (FHS) Offspring and Third Generation cohorts, who underwent proteomic profiling with SomaScan version 1.3K, we evaluated the associations between individual proteins and DTI measures. Proteins with p-values < 0.01 for associations with either FW or PSMD in the FHS cohort (exploratory phase) were selected for replication in the Age, Gene/Environment Susceptibility (AGES) - Reykjavik Study. AGES participants were profiled using a custom 5K SomaScan platform. For proteins that replicated, two-sample Mendelian randomization was conducted to explore potential causal relationships with neuroimaging traits, global cognitive function, stroke, and AD.

**Results:** A total of 48 proteins were associated with either FW or PSMD in FHS (N=1,106, 56% female, mean age=48 at protein profiling) at an uncorrected p-value < 0.01. Of these, six proteins were validated in AGES (N=2,586, 58% female, mean age=76) at p-value < 0.05/48: TFF3 (beta=0.015), LG3BP (beta=0.0089), and heparin cofactor II (HCII, beta=0.0097) showed positive associations with FW, while CCL28 was positively associated with PSMD (beta=0.01). Conversely, BMPR1A (beta=-0.016) and gelsolin (beta=-0.011) were negatively associated with PSMD, all with SE=0.002. Mendelian randomization analyses further suggested potential causal effects (p-value<0.05), including positive effects of HCII, LG3BP, and TFF3 on WMH; HCII on small vessel stroke; LG3BP on any stroke and ischemic stroke; and a negative effect of gelsolin on large artery stroke.

**Conclusions and Relevance:** This cohort and Mendelian randomization study identified several proteins significantly associated with FW and PSMD, which are potential imaging biomarkers of ischemia, neuroinflammation, and axonal degeneration. Mendelian randomization analyses suggested potential causal effects of certain proteins on white matter hyperintensities and stroke, highlighting the need for functional studies to explore whether elevated protein levels represent protective mechanisms or contribute to brain injury.

## Introduction

Alzheimer’s disease (AD) and related dementias (ADRD) affect approximately 55 million people worldwide, [1] yet safe and effective disease-modifying treatments are still lacking.[2] While abnormal protein deposition, namely extracellular β-amyloid (Aβ) plaques and intracellular tau neurofibrillary tangles (NFTs) are well-recognized pathological hallmarks of AD, [3] the cause of AD is believed to be multifactorial, and the exact pathogenesis of AD remains poorly understood. In addition to amyloid and tau, data from genome-wide association studies (GWAS) of AD also implicate important roles of proteins involved in inflammation, vascular disease, lipid processing, and endothelial injury.[4]

Diffusion tensor imaging (DTI) is a magnetic resonance imaging (MRI) technique developed in the 1990s that measures the diffusion of water molecules. DTI is increasingly used to assess white matter damage in small vessel stroke (SVS), given its sensitivity in detecting abnormalities in both white matter hyperintensities (WM) visible on T2-weighted sequences, as well as microstructural abnormalities in white matter that appeared normal on conventional MRI.[5] In recent years. two promising neuroimaging biomarkers for measuring cerebral white matter microstructural disruption have been proposed: peak width of skeletonized mean diffusivity (PSMD) and free water (FW). PSMD measures mean diffusivity (MD) along the main fiber tracts and can characterize subtle injury in white matter. Studies indicate that PSMD outperforms traditional SVD markers (e.g. white matter hyperintensity volume and lacunes) in predicting cognitive performance,[6] and is noted to be elevated in patients with cerebral amyloid angiopathy (CAA), mild cognitive impairment (MCI), and AD; as well as associated with lower processing speed domain scores in patients with CAA.[7] FW index measures the fraction of the diffusion signal explained by isotropically unrestricted water, i.e. extracellular free water and water within the vicinity of cellular tissue. FW is considered a sensitive marker of ischemia, neuroinflammation, and axonal degeneration, all important pathways in the pathogenesis of MCI and AD.[8] Elevated hippocampal free water has been reported in individuals with early MCI compared to cognitively healthy controls.[9] Higher FW has also been correlated with a higher baseline Clinical Dementia Rating (CDR) scale score and a higher risk of progression along the CDR scale.[10] In patients with MCI and AD dementia, widespread increased FW is associated with poorer attention, executive functioning, visual construction, and motor performance.[11] Thus, both PSMD and FW offer utility as early-stage non-invasive neuroimaging markers of brain injury, particularly due to AD and vascular pathology.

Association analysis of large-scale, high-throughput DNA aptamer-based plasma proteomic profiling is a promising approach to identifying novel blood biomarkers that can further our understanding of pathways underlying the pathophysiology of dementia.[12, 13] The use of neuroimaging markers of subcortical vascular disease (i.e. intermediary endophenotypes for preclinical AD) also provides the opportunity to understand the biological basis in early dementia stages. In this study, we aimed to determine the association of a large panel of circulating proteomic markers with brain imaging endophenotypes of cerebrovascular dysfunction, FW, and PSMD in community-based samples of mostly cognitively healthy adults.

## Methods

### Study Sample

#### Framingham Heart Study (FHS)

The discovery study sample consisted of mostly white participants from the Framingham Heart Study (FHS). The design of FHS has been detailed previously [14–16] and also described in the Supplementary Methods. The subjects used in this study were from the Offspring and Third generation (Gen 3) cohorts. SomaScan proteomic profiling was performed on 1,913 participants from the Offspring cohort using plasma samples collected during the fifth examination cycle (1991-1995) and on 900 participants from the Gen 3 cohort using plasma samples collected during the second examination cycle (2008-2011). DTI was performed on 1,134 Offspring participants and 2,062 Gen 3 participants during 2009-2016. After excluding participants with stroke and other neurological diseases (e.g. brain tumor, severe head trauma, Parkinson’s disease, etc.) at the time of the plasma sample collection, the final study sample consisted of 486 Offspring and 620 Gen 3 cohort participants with both protein profiling and MRI brain data. Figure 1 shows the process to obtain the final samples for association analyses. Participants all provided written informed consent. The study was approved by the Institutional Review Board at Boston University Medical Center (Protocol number H-26671).

**Figure 1.**
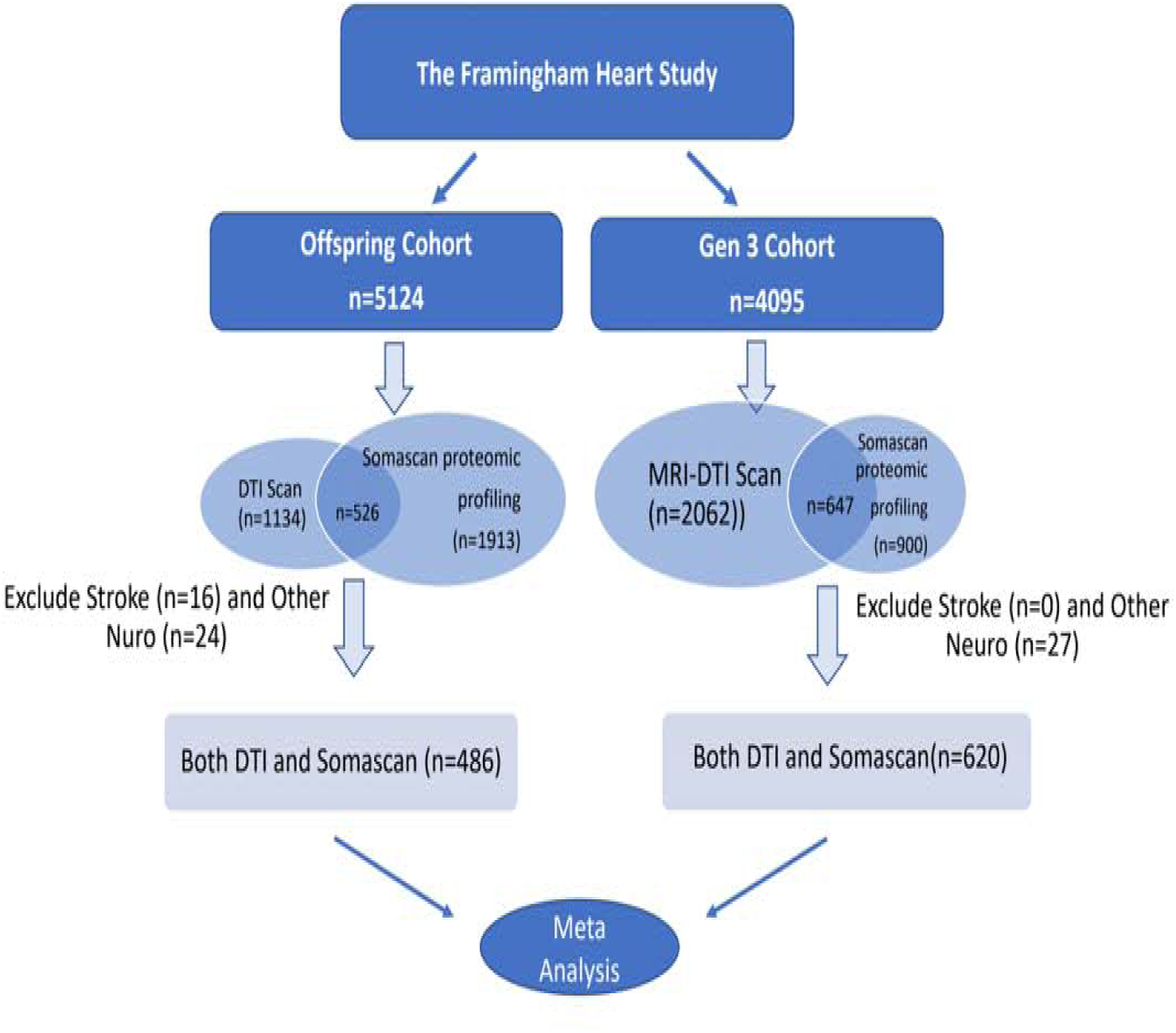
Flow-chart of obtaining the FHS samples for statistical analyses.

#### Age, Gene/Environment Susceptibility – Reykjavik Study (AGES)

The replication samples are from AGES which was initiated in 2002. AGES is a single-center prospective population-based study of deeply phenotyped subjects (n = 5,764, mean age 76.6±5.6 years) and survivors of the 40-year-long prospective Reykjavik study, an epidemiologic study aimed to understand aging in the context of gene/environment interaction.[17] Of 5,457 individuals with protein measurements, 2,586 with PSMD and FW measurements after applying the same exclusion criteria as FHS (n exclusion =267) were included in the replication analyses. The study was approved by the Icelandic National Bioethics Committee (VSN: 00-063).

### FW and PSMD Measurement

In FHS, FW and PSMD were computed from MRI performed using a 1.5-Tesla Siemens Avanto scanner, with a diffusion tensor imaging (DTI) sequence. [18] Specifically, DTI was performed using the following parameters: repetition time (TR)=3600 ms, echo time (TE)=94 ms, 25 slices total, field of view=25 cm, acquisition matrix =128 × 128, slice thickness = 5 mm with 5 mm gap. Diffusion weighted images were generated using 30 gradients directions with total gradient diffusion sensitivity of b=1000 s/mm2, and one image with b =0 s/mm2. Centralized reading of all images was performed using in-house designed imaging, visualization and analysis software (Quanta 2). FW measures water molecules that do not experience flow and are not restricted by their surroundings. A free water compartment, which models isotropic diffusion with a diffusion coefficient of water at body temperature fixed to 3 × 10−3 mm2/s, and a tissue compartment, which accounts for all intra and extracellular molecules that are hindered or restricted by tissue membranes were modeled.[8] The fractional volume of the free water compartment in each voxel was estimated, resulting in the FW map. A white matter (WM) mask was defined by thresholding the FSL(Functional Magnetic Resonance Imaging of the Brain @ Oxford Software Library)[19] FA template at a value of 0.3 to reduce CSF partial volume contamination. For each individual, an overall measure of mean FW was computed by superimposing the WM mask onto the respective individual coregistered FW maps and averaging FW values within these WM voxels. PSMD was designed to eliminate the contaminating signal of cerebrospinal fluid from DTI-derived MD maps. It was calculated following an automatic procedure [20] using the freely available scripts (http://www.psmd-marker.com). Briefly, the DT-MRI data were processed using the standard Tract-based Spatial Statistics (TBSS)[21] pipeline available in FSL with histogram analysis performed on the resulting white matter MD skeletons. PSMD was then calculated as the difference between the 95th and 5th percentiles of the voxel-based MD values within each participant’s white matter tract skeleton. The PSMD and FW measurements in AGES followed the same protocol.

### Proteomic Profiling

At the baseline visit of FHS (examination cycle 5 for the Offspring cohort and examination cycle 2 for the Gen 3 cohort), fasting blood samples were obtained from the antecubital vein of participants who were placed in the supine position. Blood samples were immediately centrifuged before being stored at -80°C. Plasma protein concentrations were subsequently measured on these stored samples using SomaScan® Human Plasma 1.3K version, a single-stranded DNA aptamer-based platform. [22] For Offspring samples, the median intraassay CV was < 4% and median interassay CV was < 7% across batches. For Gen 3 samples, both median intra- and interassay CVs were < 4%.

The proteomic measurements in AGES have been described in detail elsewhere[23] and were available for 5,457 participants. Briefly, a custom version of the SOMAscan platform (Novartis V3-5K) was applied based on the slow-off rate modified aptamer (SOMAmer) protein profiling technology including 4,782 aptamers that bind to 4,137 human proteins. Serum was prepared using a standardized protocol from blood samples collected after an overnight fast, stored in 0.51ml aliquots at −80°C, and serum samples that had not been previously thawed were used for the protein measurements. All samples were run as a single set at SomaLogic Inc. (Boulder, CO, US).

### Protein-MRI Phenotype Association Analyses in Discovery Cohort

Prior to association analyses, the phenotypes FW and PSMD were natural logarithm-transformed to approximate a normal distribution. To test protein-phenotype associations in FHS, we fit linear mixed effects models for Offspring and Gen 3 separately, with each phenotype as the dependent variable and each log-standardized protein as an independent variable, adjusting for age at MRI, sex, total intracranial volume, time difference between blood draw and completion of MRI, blood sample batch, and plate. Inverse-variance weighted meta-analyses were performed to combine FHS cohort-specific results of each phenotype-protein association analysis. For top associated proteins in FHS, we queried the published data on correlation between SomaScan and Olink measures of 1,069 proteins estimated using 485 individuals from the Fineland study [24] to confirm the specificity of the aptamers.

### Protein-MRI Phenotype Association Analyses in Replication Cohort and Meta Analyses

We identified proteins with association p-values < 0.01 with either FW and PSMD in FHS for replication in AGES using a linear regression model, adjusting for age, sex, and total intracranial volume. This moderately stringent threshold was chosen to balance the discovery sensitivity with false positive control, consistent with the hypothesis-generating design of this initial discovery phase. Inverse-variance weighted fixed effects meta-analyses were performed to combine the FHS and AGES association results for the selected proteins with FW and PSMD.

### Mendelian Randomization Analyses

For proteins that were replicated in AGES, we performed two-sample Mendelian randomization studies [25] using previously reported protein quantitative trait loci (PQTLs) as instrumental variables (IVs). These pQTLs were single nucleotide polymorphisms (SNPs) identified from previous GWAS of the proteins (exposures) with genome-wide significance and with no or very low linkage disequilibrium between each other.

Both cis- and trans-pQTLs were included as instrumental variables (IVs) in the initial analyses. However, additional analyses were conducted using only cis-pQTLs (when applicable, defined as variants within ±500 kb of the protein-encoding genes) as IVs, as these are less likely to exhibit horizontal pleiotropy compared to trans-pQTLs.

We tested the putative causal relationships between exposure and *apriori* selected outcomes including AD, MRI, cognitive and stroke phenotypes in the Mendelian randomization analyses. To obtain pQTL-outcomes association results, we leveraged previous GWAS for white matter hyperintensity (WMH),[29] AD, [30] general cognitive function,[31] and stroke phenotypes, including any stroke (AS), ischemic stroke (IS), and ischemic stroke subtypes (LAS -large artery stroke, SVS – small vessel stroke, and CES – cardioembolic stroke ).[32] Since the alleles at genetic loci are randomly inherited from parents, uninfluenced by environmental factors, when there are no other paths for the instrumental variants to affect an outcome bypassing the exposure (i.e. no horizontal pleiotropy), the association of a genetic exposure with an outcome indicates a causal relationship.[33] The inverse-variance weighted (IVW) method was used to combine the evidence based on multiple genetic instruments.[25] For protein-outcome pairs with a significant IVW test (p-value<0.05) in our primary analyses, we additionally applied pleiotropy-robust sensitivity analyses, including weighted median, weighted mode, and MR Egger where applicable.[34]

### Colocalization Analyses

We applied the approximate Bayes factor method[35] to assess whether the exposures (proteins) and outcomes share the same causal variant in the region of a cis-pQTL. For each lead cis-pQTL defined by having the lowest p-value among all p-QTLs within ±200 kilobase(kb) distance, we included all SNPs within 200kb region of the lead pQTL in the analyses. Five mutually exclusive hypotheses were evaluated within the region: (H0): no causal variant to the protein or the outcome, (H1) only one causal variant for the protein, (H2) only one causal variant for the outcome, (H3) two distinct causal variants for the protein and outcome, and (H4) protein and outcome sharing a causal variant, each using a posterior probability, PP0 – PP4 respectively. We used default conservative priors *p*1= 10^−4^, *p*2= 10^−4^ and *p*12 = 10^−5^, where *p*1 and *p*2 are the probabilities that a given variant is associated with either protein or outcome and *p*12 is the probability that a given variant is associated with both traits. We use the conventional threshold of PP4≥0.75 as evidence of colocalization, i.e. the two traits share the same causal variant.

### Protein-Protein Interaction Network Analyses

Protein-protein interaction (PPI) network analyses were performed in STRING (‘Search Tool for the Retrieval of Interacting Genes/Proteins’) Version 11.5[36] using the top associated proteins (p-value <0.01) for each phenotype in FHS. Details of PPI network construction can be found in Supplementary Materials.

Enrichment analysis was also carried out for gene ontologies, pathways, and domains, which shows terms that were more enriched in the set of proteins in the network than by random.

### Pathway and Network Analyses using Ingenuity Pathway Analysis (IPA) system

We used the Core Analysis in IPA to interpret the top associated proteins (p-value <0.01) for each phenotype in the context of biological processes, pathways, and networks using literature-curated Ingenuity Knowledge Base (genes only).[37] Details can be found in Supplementary Materials.

## Results

### Sample Characteristics

Table 1 summarizes the baseline characteristics of the discovery and replication cohort samples from FHS and AGES. In the FHS Offspring cohort, 58% of participants were women, with a mean age of 51 years at blood collection and 71 years at brain MRI. In the FHS Gen 3 cohort, 54% of participants were women, with a mean age of 46 years at blood collection and 47 years at brain MRI. In the AGES cohort, 58% of participants were women, and the mean age was 76 years for both blood collection and brain MRI.

**Table 1.** Characteristics of the discovery and replication samples

|  | FHS |  |  |  | AGES |  |
| --- | --- | --- | --- | --- | --- | --- |
|  | Offspring (N = 486) |  | Gen3 (N = 620) |  | (N = 2586) |  |
|  | mean | SD | mean | SD | mean | SD |
| female(%N) | 58 (280) |  | 54 (336) |  | 58 (1494) |  |
| age at MRI | 71.40 | 7.98 | 47.21 | 8.12 | 76.40 | 5.31 |
| age at protein profiling | 50.47 | 8.07 | 45.46 | 8.01 | 76.40 | 5.31 |
| age difference (MRI vs. protein profiling) | 20.93 | 1.49 | 1.75 | 1.03 | 0.00 | 0.00 |
| total cranial volume (mL) | 1269.64 | 131.95 | 1293.25 | 130.20 | 1496.29 | 146.27 |
| free water | 0.23 | 0.03 | 0.20 | 0.02 | 0.19 | 0.032 |
| log(free water) | -1.46 | 0.14 | -1.63 | 0.08 | -1.69 | 0.16 |
| PSMD | 2.8E-04 | 5.7E-05 | 2.1E-04 | 2.5E-05 | 0.00037 | 7.5E-05 |
| log(PSMD) | -8.20 | 0.19 | -8.44 | 0.11 | -7.92 | 0.17 |

### Protein-MRI Phenotype Associations

Figure 2 presents volcano plots of the protein-phenotype association results from FHS. Supplementary Table 1 provides the association results for 48 proteins (36 unique) identified in FHS as significantly associated (p < 0.01) with FW (n = 23) or PSMD (n = 25), along with their replication results in AGES and the meta-analyses results combining FHS and AGES. Replication analyses in AGES identified six proteins as significant at alpha=0.05/48. The replicated proteins were TFF3, heparin cofactor II (HCII), LG3BP, BMPR1A, gelsolin and CCL28 (results presented in Figure 3).

**Figure 2.**
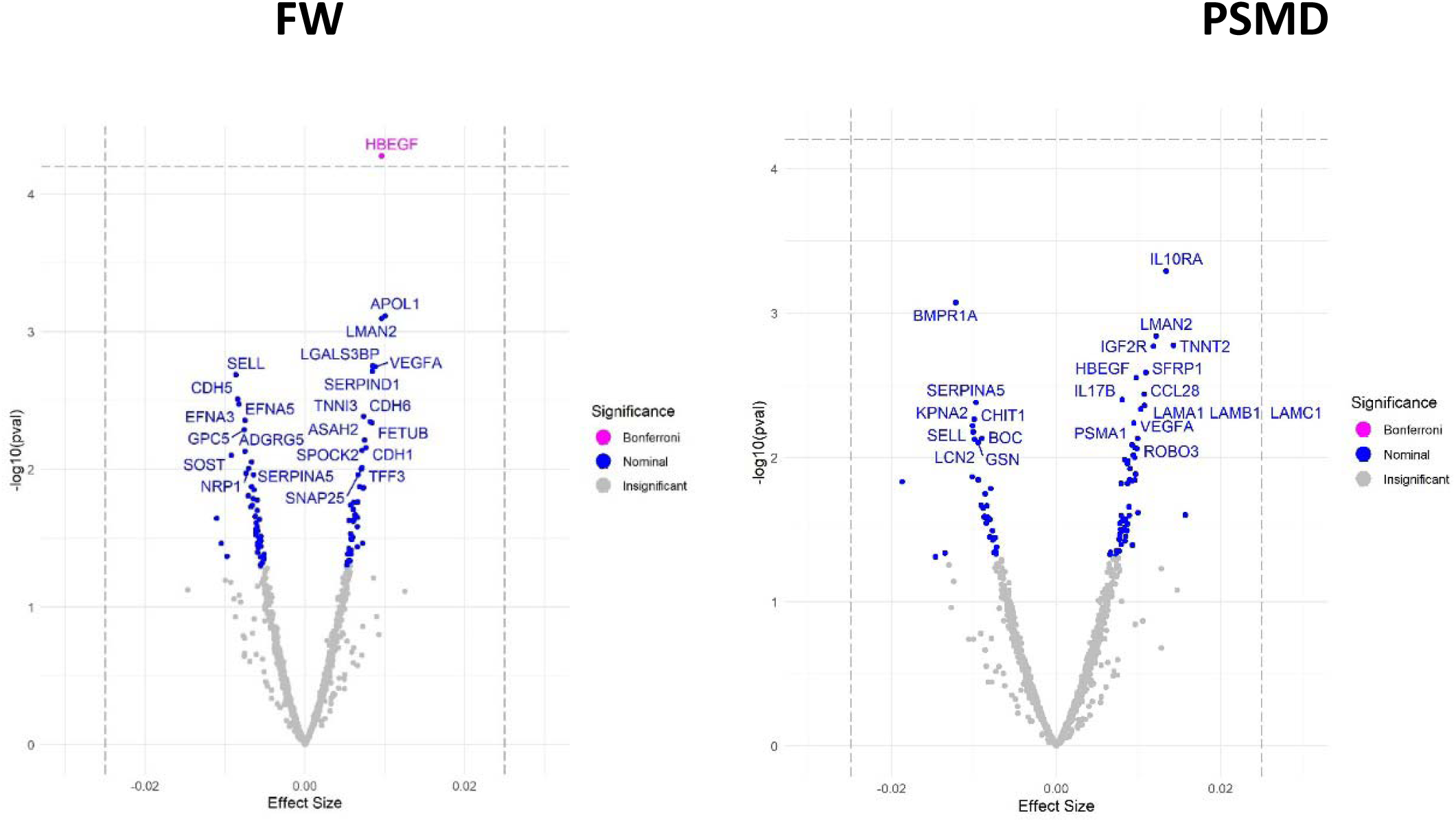
Volcano plots displaying FW and PSMD association results for all proteins in the discovery cohort. The X-axis is the size of the association effect, and the y-axis is the –log_10_ transformed p-value.

**Figure 3.**
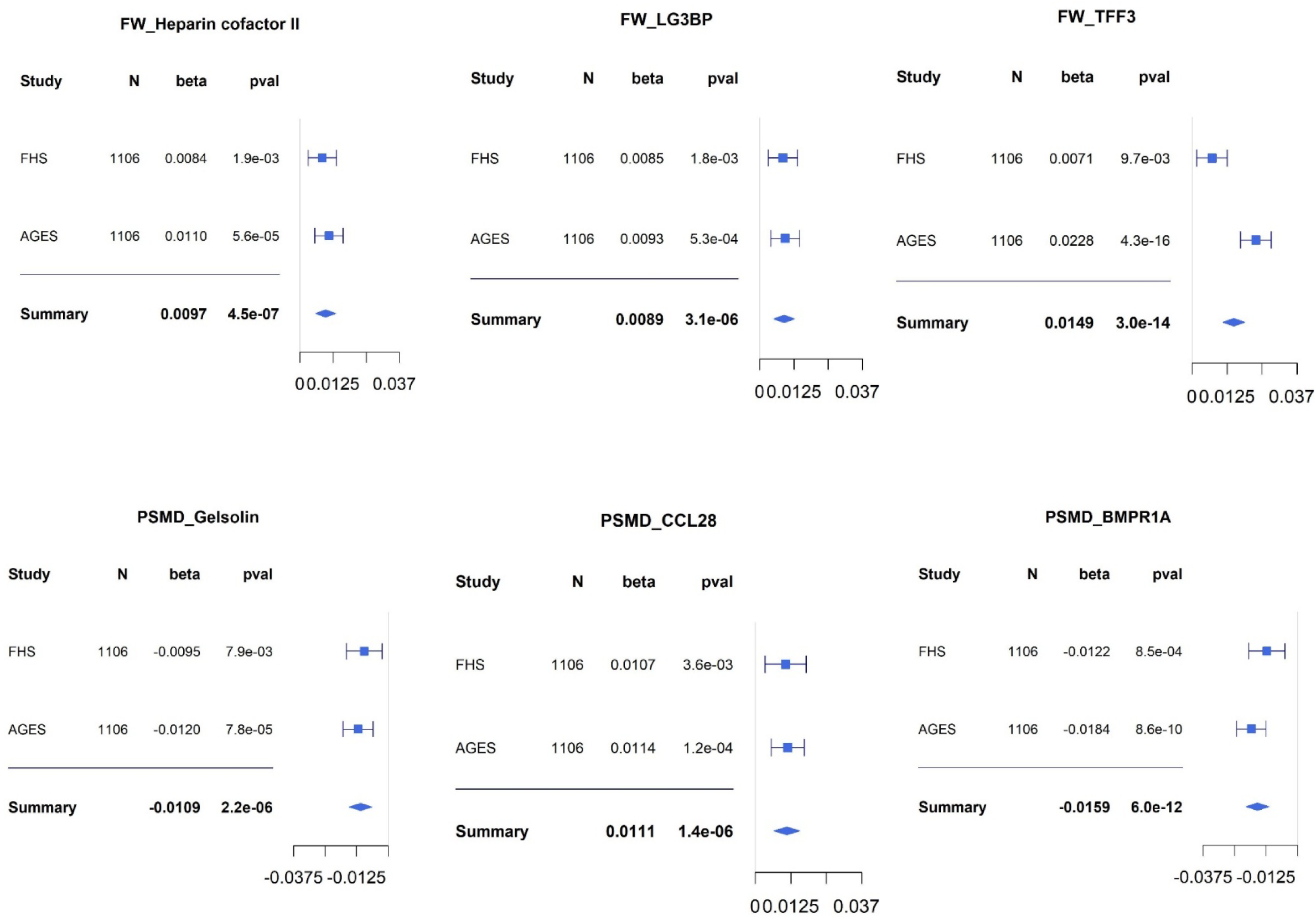
Forest plot of the protein-phenotype associations for replicated proteins in discovery and replication cohorts.

In the meta-analyses, higher TFF3 levels were significantly associated with increased FW (beta = 0.015, SE = 0.0020, p = 2.98 × 10 ¹ ), as were elevated HCII (beta = 0.0097, SE = 0.0019, p = 4.53 × 10 ) and LG3BP (beta = 0.0089, SE = 0.0019, p = 3.09 × 10 ). Increased BMPR1A (beta = -0.016, SE = 0.0023, p = 6.01 × 10 ¹²) and gelsolin (beta = -0.011, SE = 0.0023, p = 2.18 × 10 ) were associated with lower PSMD, while higher CCL28 was linked to increased PSMD (beta = 0.0111, SE = 0.0023, p = 1.41 × 10 ).

An additional 10 proteins (HB-EGF, ASAH2, BOC, Cadherin E, Cadherin-5, Cadherin-6, Gro-b/g, IGF-II receptor, Lectin; mannose-binding 2, and SARP-2) showed nominal associations (p < 0.05) with either FW or PSMD in the replication analysis (Supplementary Table 1s). However, only six associations (Cadherin E, Cadherin-5, Gro-b/g, SARP-2, IGF-II receptor, and BOC) were directionally consistent across the two studies (Supplementary Figure 1s).

The median Spearman’s correlation between SomaScan and Olink profiling was 0.74 for the top proteins associated with FW (p < 0.01, n = 14/23) and 0.68 for those associated with PSMD (n = 12/25). The 25th and 75th percentiles were [0.55, 0.78] for FW proteins and [0.35, 0.75] for PSMD proteins.

### Mendelian Randomization Analyses

Five out of the six replicated proteins (CCL28, HCII, LG3BP, TFF3, gelsolin) had pQTLs reported in existing literature,[28] with 109 pQTLs (27 in cis regions) used as instrumental variables (IVs) in the Mendelian randomization study (Supplementary Table 2s). The F-statistics for all pQTLs exceeded the threshold of F = 10, indicating robust instruments.

Mendelian randomization analysis suggested a potential causal association at uncorrected alpha=0.05 between HCII and AD (beta = 0.061, SE = 0.026, p = 0.018) when both trans- and cis-pQTLs were included as IVs, though this association was not observed when using only cis-pQTLs (Figure 4). Two pleiotropy robust methods (Weighted median and Weighted mode) showed similar effects (beta = 0.06-0.07, p<0.01), but this is not the case with the MR-Egger test (beta=-0.0097, p=0.75). Conversely, HCII was associated with WMH (beta = 0.18, SE = 0.085, p = 0.032) and SVD (beta = 0.97, SE = 0.336, p = 0.0040) when only cis-pQTLs were included, but these associations disappeared when both trans- and cis-pQTLs were included. Since the number of cis-pQTLs is less than two, we were not able to apply pleiotropy-robust methods that require three or more genetic instruments.

**Figure 4.**
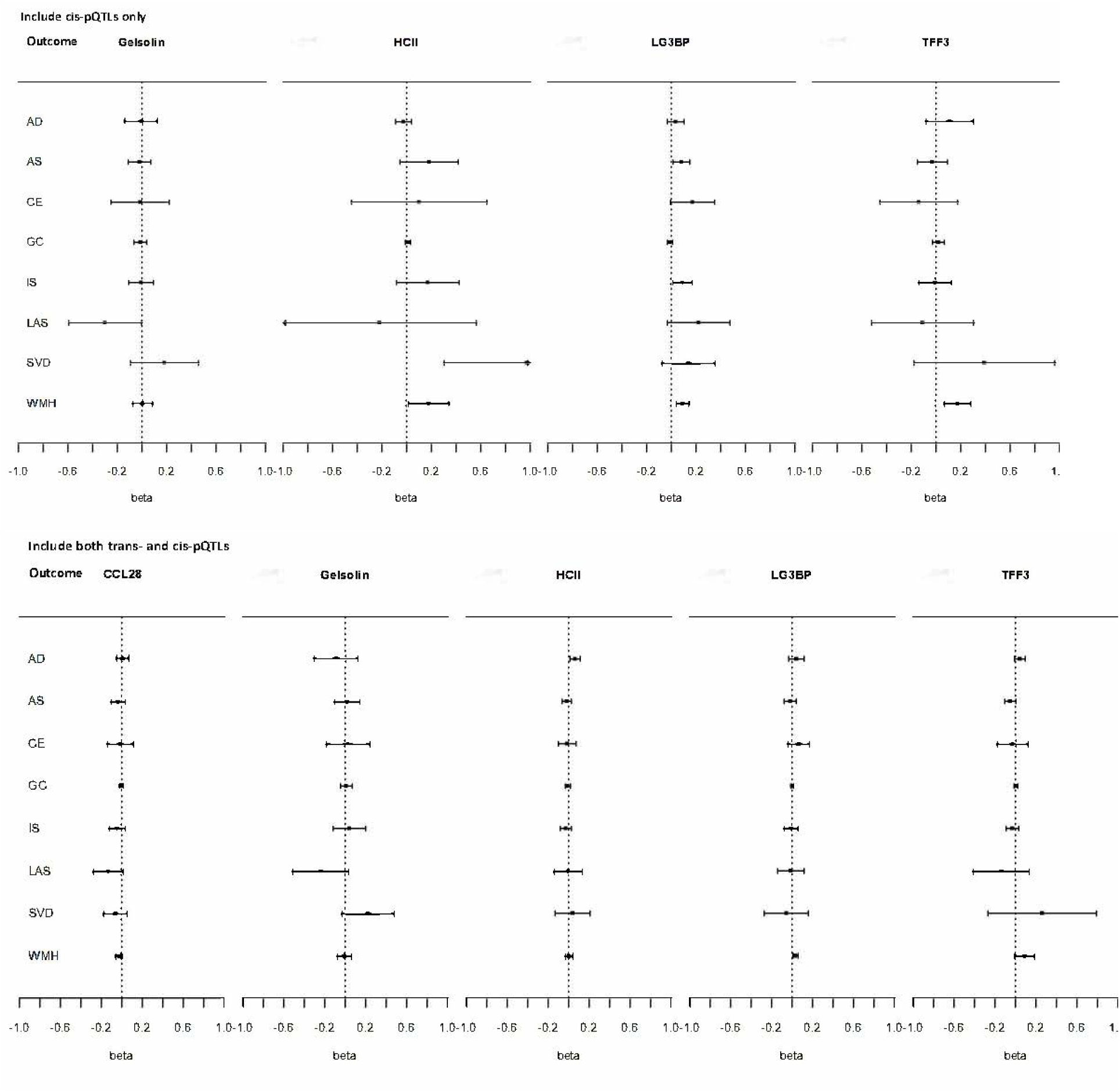
Forest plot of Mendelian randomization study results relating proteins to cognitive, MRI and stroke outcomes. X-axis is the estimated effect size of unit increase in genetically determined protein levels on the outcomes, with 95% confidence intervals. Top panel: Cis-pQTLs as instruments Bottom panel: Cis- and Trans-pQTLs as instruments

LG3BP (beta = 0.031, SE = 0.017, p = 0.015) and TFF3 (beta = 0.095, SE = 0.047, p = 0.045) were associated with WMH when both trans- and cis-pQTLs were used, with stronger associations observed when only cis-pQTLs were included (Supplementary Tables 3s, 4s). Pleiotropy robust methods yielded similar effect sizes but only the p-value of weighted median method reached nominal significance using only cis-pQTL. Using only cis-pQTLs, LG3BP was associated with AS (beta = 0.08, SE = 0.035, p = 0.022) and IS (beta = 0.09, SE = 0.0395, p = 0.021), while gelsolin appeared protective against LAS (beta = -0.30, SE = 0.15, p = 0.047); these associations were not significant when including both trans- and cis-pQTLs as IVs. (Supplementary Tables 3s, 4s). We were not able to apply pleiotropy-robust methods to validate cis-pQTL-only results due to an insufficient number of cis-pQTLs.

### Colocalization Analyses

Colocalization analyese results are presented in supplementary Table 4s. There was strong evidence that Gelsolin shared the same causal variant with AS (PP4=0.98) and IS (PP4=0.90), but weak evidence for a common causal variant between HCII and SVD (PP4=0.33), and between TFF3 and SVD (PP4=0.25); little evidence of distinct causal variants (PP3≤0.03 for all). For all other protein-outcome pairsPP4<0.1 and PP3 <0.17

### Protein-Protein Interaction (PPI) Network

The PPI network has significantly more interactions than expected (16 fitted vs. 5 expected edges, p-value=0.00016 for FW; 20 fitted vs. 7 expected edges, p-value=1.98x10^-5^ for PSMD). Vascular endothelial growth factor-A (VEGFA) had the most edges and was involved in 3 out of the top 4 significant Gene Ontology terms as defined by strength for molecular function enrichment. Additional enrichment results are presented in Supplementary Table 5s-6s and interaction networks are presented in Supplementary Figure 2s-3s.

### Pathway and Network Analyses using IPA

Supplementary Tables 7s and 8s present IPA pathway and network analysis results for FW and PSMD, respectively. The top Ingenuity Canonical Pathway was the synaptogenesis signaling pathway. The top upstream regulator was the estrogen receptor. For downstream functions, the top diseases and disorders included cardiovascular disease, hematological disease, and organismal injury and abnormalities. The top two networks are presented in Supplementary Figure 4s-5s for FW, and Supplementary Figure 6s-7s for PSMD.

## Discussion

We identified six proteins associated with FW or PSMD across two independent cohorts, FHS and AGES. The identified proteins were TFF3, LG3BP, HCII, BMPR1A, CCL28 and gesolin. LG3BP has recently been shown to play a role in human cortical complexity [38] and was found elevated in CSF and associated with Aβ deposits and tau aggregates in brain tissue in Alzheimer’s disease.[39] BMPR1A is a member of the bone morphogenetic protein (BMP) receptor family, which plays a role in embryogenesis and development of multiple organ systems, including the vasculature and central nervous system, and was found protective against incident heart failure [40]. *BMPR1A* gene was associated with AD and a potential drug target whose expression levels in the entorhinal cortex were lower in AD than in control samples. [40] [41] In animal models, the expression of *BMPR1* was strongly decreased in AD kidneys but recovered after increased training [42]. HCII, heparin cofactor II, is a serine protease that inhibits thrombin. HCII has been proposed to regulate coagulation or to participate in processes such as inflammation, atherosclerosis, and wound repair.[43] HCII was found to increase in early and later stages of autosomal dominant Alzheimer’s disease compared to normal noncarrier control and inversely associated with amyloid deposition in the caudate nucleus. [44] In our Mendelian Randomization study, HCII was significantly associated with an increased risk of small vessel stroke, raising the possibility of HCII playing a role in AD via vascular pathways. CCL28 is a mucosal chemokine with antimicrobial properties. Compared with control mice, CCL28 was upregulated in the brain of APP/PS1 mice of Alzheimer’s disease. In our Mendelian Randomization study, increased CCL28 was borderline significantly associated with higher white matter hyperintensity. TFF3, intestinal trefoil factor (TFF3 or ITF), is one of the three trefoil factor peptides (TFFs) secreted by goblet cells and epithelial cells of various tissues in humans. Recent studies suggested that TFF3 was present not only in the intestinal tract but also in other organs including brain.[45] Among a group of cerebrospinal fluid (CSF) biomarkers, TFF3 is the strongest predictor of brain atrophy, with lower levels of TFF3 in CSF associated with greater brain atrophy in individuals with AD pathology. [46] Our Mendelian randomization study found lower levels of plasma TFF3 associated with lower FW but higher WMH. Gelsolin, a multifunctional actin-binding protein, binds Aβ, inhibits its aggregation into fibrils, and protects cells from apoptosis induced by Aβ. Furthermore, administration of gelsolin significantly reduced amyloid load and Aβ level in AD transgenic mice.[47] These biomarkers collectively highlight pathways of inflammation, vascular dysfunction, and tissue repair implicated in ADRD.

Strengths of our study include the availability of over 1,300 proteins in a large prospective community-based cohort, FHS, allowing us to identify proteins potentially implicated across diverse pathophysiological pathways of AD dementia. We evaluated associations with two novel neuroimaging endophenotypes which are highly sensitive to subtle preclinical white matter changes. Thus, we were able to evaluate biomarker associations with the earliest signs of white matter injury many years prior to detectable changes on conventional MRI sequences. We validated some of our top findings in another cohort, AGES, greatly decreasing the likelihood of false positive findings for replicated associations. However, our study also has a number of limitations. While FW and PSMD provide sensitive measures of global white matter integrity, they lack topographic specificity. Future studies incorporating tractography or atlas-based segmentation could delineate which specific white matter tracts (e.g., corpus callosum, superior longitudinal fasciculus) are most strongly associated with proteomic alterations, offering deeper mechanistic insights. The mean age at blood sample collection was 20 years younger for FHS samples compared to AGEs samples. However, the FHS offspring cohort and AGEs sample have similar mean ages at DTI imaging. Because of this, this study was more likely to detect proteins with both diagnostic and prognostic association with the DTI phenotypes and less likely to detect proteins with only one type of association, thus less likely to be subject to bias than a cross-sectional study. The number of cis-pQTL for each protein is quite small (n=1 or 2, except for LG3BP), making it impossible to assess pleiotropy statistically, which requires three or more instruments. While cis-pQTLs are less prone to horizontal pleiotropy than trans-pQTLs, some cis variants can influence expression levels of non-cognate genes and hence proteins encoded by these genes. One cis-pQTL of HCII and two cis-pQTLs of gelsolin were also pQTLs of other proteins, but the cis-pQTLs of the rest of the proteins were not significantly associated with other proteins. LG3BP’s associations with AS and WMH identified using IVW where there are >2 genetic instruments were also supported by pleiotropy robust weighted median test but not by the MR-egger test; however, the latter was well known for limited power. Our colocalization analyses results only provided weak evidence of the same causal variant between HCII and SVD, raising the likelihood of MR assumption violation: the IV may affect SVD indirectly through the proteins. Further analyses and biological experiments are warranted to pinpoint the exact roles of these proteins in AD pathology. Lastly, our study participants are primarily Caucasian, thus our findings may not be generalizable to other ethnicities.

In summary, our study has identified several proteins associated with free water and PSMD, neuroimaging markers of ischemia, neuroinflammation, and axonal degeneration implicated in many neurological diseases, including mild cognitive impairment and Alzheimer’s disease. Pathway and network analyses of top proteins suggest pathways of cell-to-cell signal, adhesion and inflammation mediation for the discovered proteins. Further observational and experimental studies are warranted to understand better the biological mechanism of these proteins and their potential as a treatment target for AD.

## Data Availability

All data produced in the present study are available upon reasonable request to the authors

## Acknowledgment

FHS: The Framingham Heart Study is conducted and supported by the National Heart, Lung, and Blood Institute (NHLBI) in collaboration with Boston University (Contract No. N01-HC-25195, HHSN268201500001I and 75N92019D00031). This work was also supported by grant R01AG063507, R01AG054076, R01AG049607, R01AG059421, R01AG033040, R01AG066524, P30AG066546, U01 AG052409, U01 AG058589 from the National Institute on Aging and R01 AG017950, UH2/3 NS100605, UF1 NS125513 from National Institute of Neurological Disorders and Stroke to S.S and R01HL132320 from National Heart, Lung, and Blood Institute to R.E.G., V.R., and S.S.

AGES: The Age Gene/Environment Susceptibility – Reykjavik Study has been funded by NIA contracts N01-AG012100 and HSSN271201200022C, NIH Grant No. 1R01AG065596-01A1, Hjartavernd (the Icelandic Heart Association), and the Althingi (the Icelandic Parliament).

