## Supplementary Materials for "Plasma Proteomics and Sensitive Imaging Biomarkers of Vascular Brain Injury"

for

### **Supplementary Methods**

#### ***Discovery Study Sample of FHS***

The study sample consisted of participants from the Framingham Heart Study (FHS). The design of FHS has been detailed previously.[1-3] In brief, the study was initiated in 1948 with an enrollment of 5209 mostly Caucasian individuals from the town of Framingham, MA, USA. This cohort is referred to as the Original cohort. Starting in 1971, the offspring of the Original Cohort along with their spouses (n=5124), were enrolled in the ‘Offspring cohort’ and have been examined at 4-yearly intervals thereafter. In 2002, the children of the Offspring cohort were enrolled as the ‘Third generation cohort’ (Gen 3) and are examined periodically with a target interval of 4 years between examinations, although for logistic reasons some exams were 6-8 years apart (n=4095). SomaScan proteomic profiling were performed on 1,913 participants from the Offspring cohort using plasma samples collected during the fifth examination cycle (1991-1995) and 900 participants from Gen 3 cohort using plasma samples collected during the second examination cycle (2008-2011).

An optional MRI brain scan was offered to all Offspring participants who attended examination cycle 8 and to Gen 3 cohort participants who attended examination cycle 2. A total of 1,134 Offspring participants and 2,062 Gen 3 particiapnts had MRI-DTI performed during 2009-2016. For the purposes of this investigation, we excluded 23 Offspring and 12 Gen 3 cohort particpants with stroke and an additional 53 Offspring and 74 Gen 3 participants with other neurological diseases (e.g. brain tumor, severe head trauma, Parkinson’s disease, etc) at the time of the plasma sample collection. The final study sample consisted of 486 Offspring and 620 Gen 3 cohort participants with both protein profiling and MRI brain data. Association analyses were performed in each cohort separately then combined using meta analyses as detailed below in Protein-MRI Phenotype Association Analyses. Figure 1 shows the process to obtain the final samples for association analyses. Participants had all provided written informed consent. The study was approved by the Institutional Review Board at Boston University Medical Center.

#### ***Protein-Protein Interaction Network Analyses***

Protein-protein interaction (PPI) network analyses were performed in STRING (‘Search Tool for the Retrieval of Interacting Genes/Proteins’) Version 11.5[9] using the proteins demonstrating the most significant associations (p-value <0.01) for each individual phenotype. A PPI network is constructed by coding genes of input proteins as nodes. Two proteins are connected by an edge if they are associated functionally or physically with a confidence score higher than the cutoff for medium confidence (0.4). Confidence scores can be imported from curated or experimental databases (e.g. BIND, HPRD, MINT, BioCyc, GO, KEGG, Reactome), predicted by STRING based on systematic genome comparisons[10] or derived by benchmarking the performance of the predictions against a common reference set of trusted, true associations (e.g. KEGG), in addition to text mining, co-expression, and protein homology. The number of edges in the network is then compared to the number expected, presuming nodes were randomly selected from a database of all proteins. A PPI enrichment p-value is the proportion of networks of randomly selected proteins where the number of edges exceeds that of the input protein network.

K-means clustering was performed using the distance matrix obtained from the STRING global scores, such that proteins with a higher global score had a greater chance of ending up in the same cluster

#### ***Pathway and Network Analyses using Ingenuity Pathway Analysis (IPA) system***

We used the Core Analysis in IPA to interpret the top associated proteins (p-value <0.01) for each phenotype in the context of biological processes, pathways, and networks using literature curated Ingenuity Knowledge Base (genes only).[11] Proteins were overlaid onto global molecular networks developed from information contained in the knowledge base in IPA. IPA then performed: i) Canonical Pathways Analysis to identify function-specific genes significantly present within the generated networks; ii) Upstream Regulator Analysis (URA) to determine likely upstream regulators directly or indirectly connected to coding genes; iii) Downstream Effects Analysis to identify diseases, functions, and toxicity that are expected to increase or decrease, given the observed protein effects in our dataset. Downstream Effects Analysis is based on expected causal effects between molecules and functions; (iv) Causal Network Analysis (CNA) as a generalization of URA including paths that involve more than one link (i.e. through intermediate regulators) to generate a more complete picture of possible root causes for the effects of the proteins; (v) Regulator Effect Networks that connect upstream regulators, dataset molecules and downstream functions to explain how the phenotype is regulated by activated or inhibited upstream regulators.

### **Supplementary Figures**

**Figure 1s.** Forrest plots of non-replicated proteins but with a nominally significant association p-value with FW or PSMD in replication cohort.

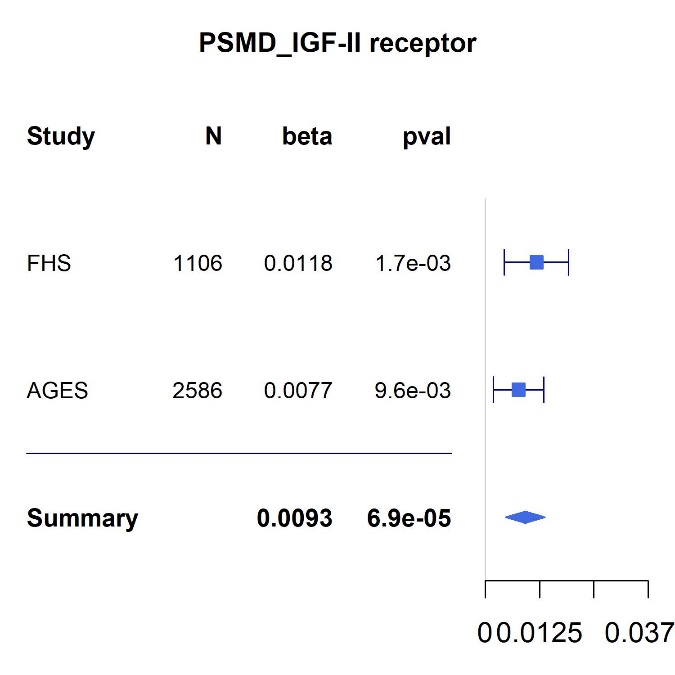

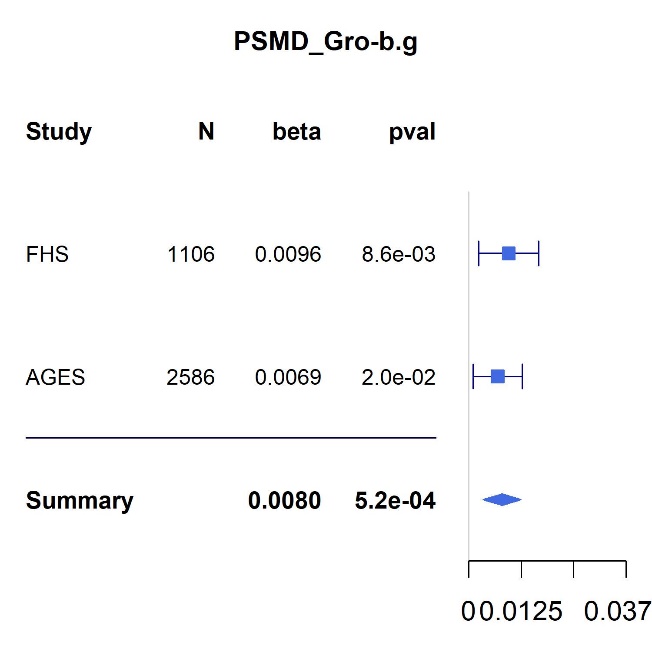

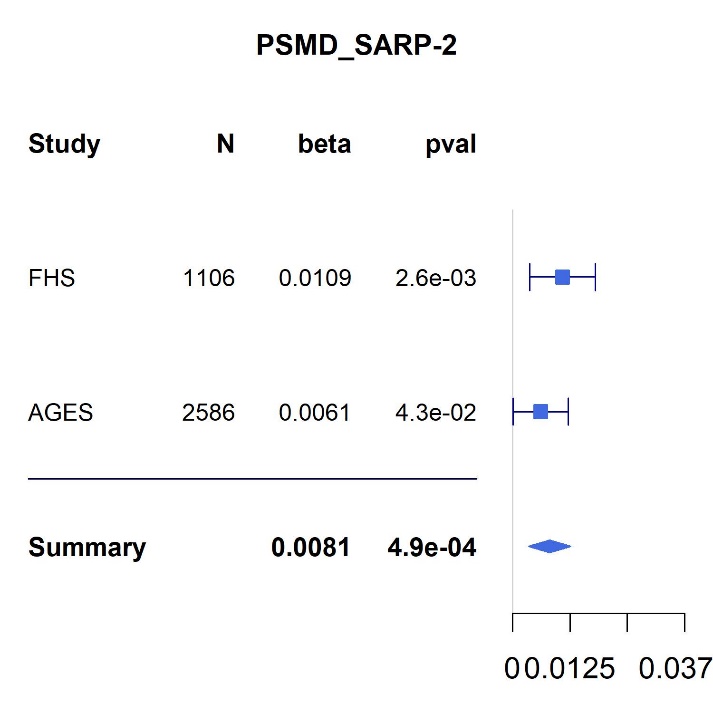

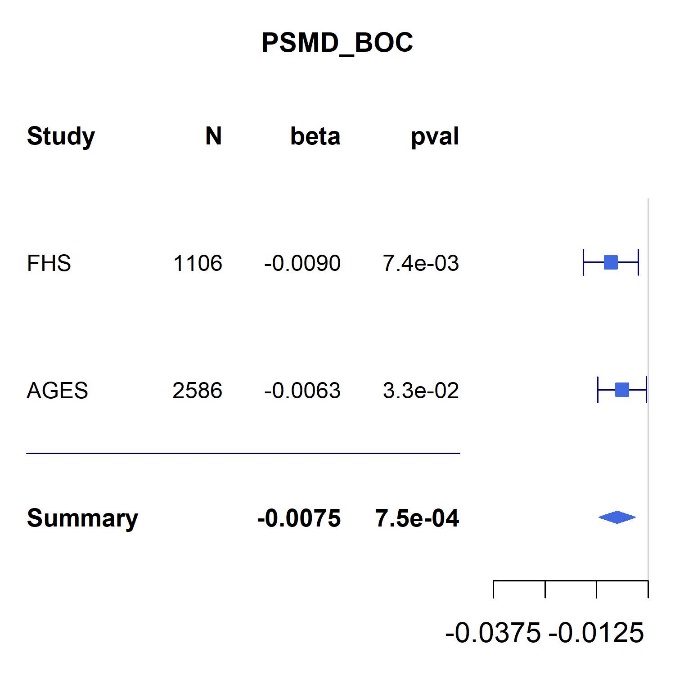

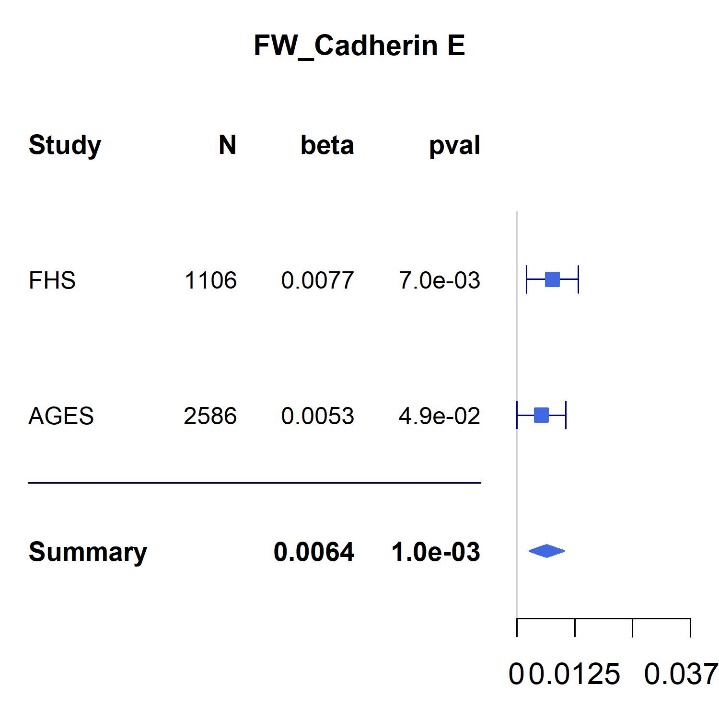

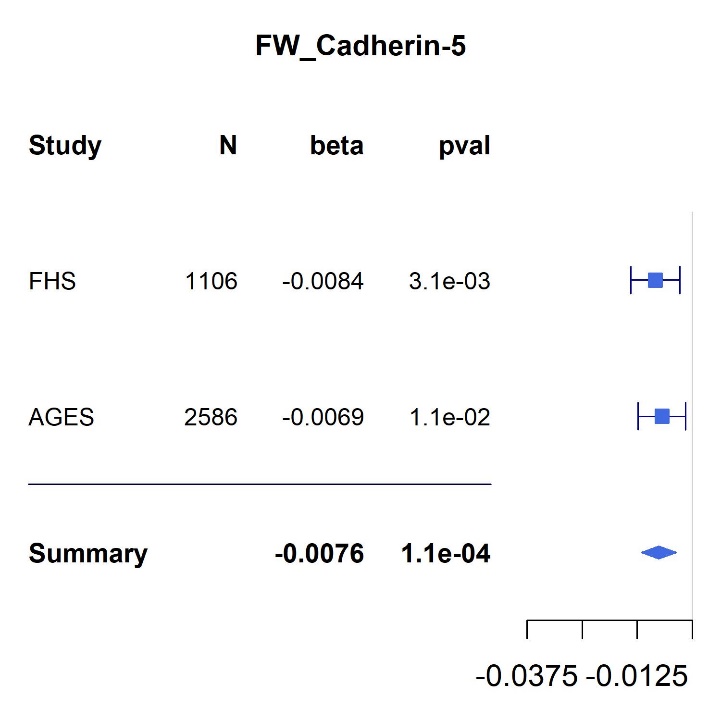

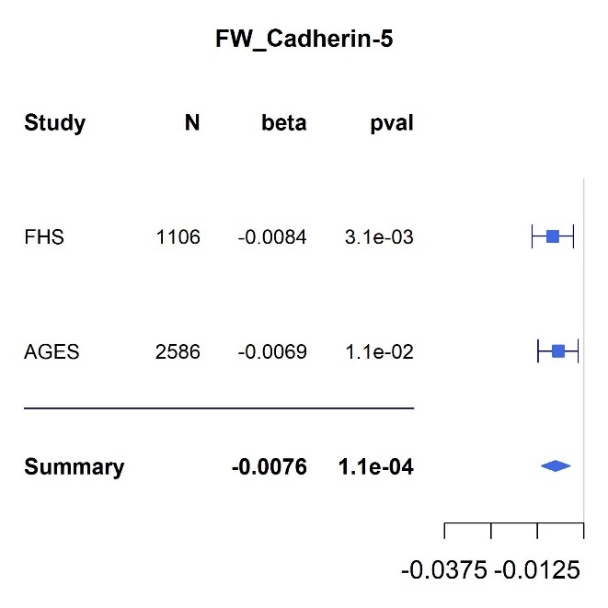

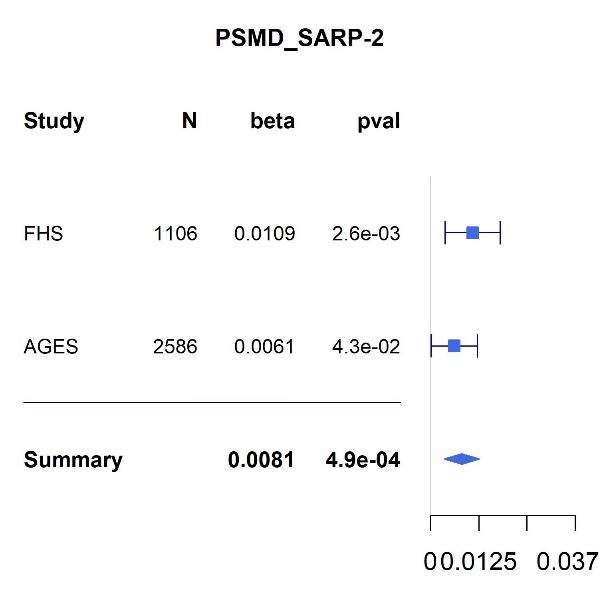

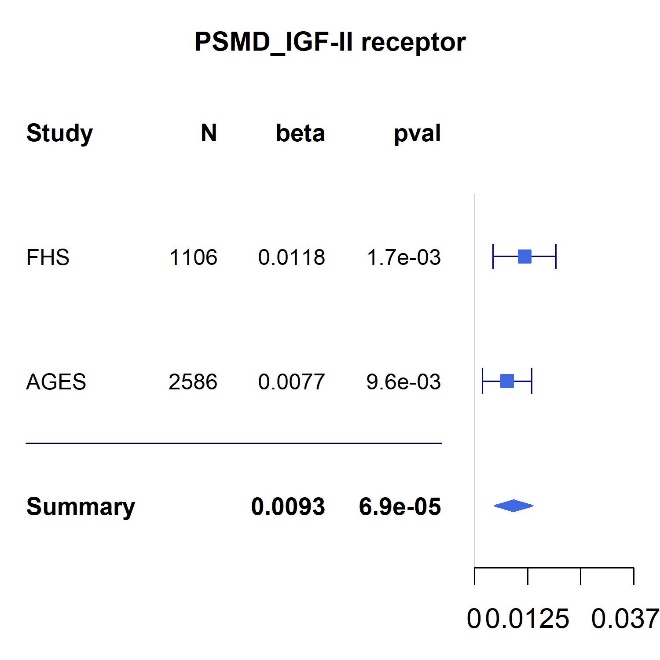

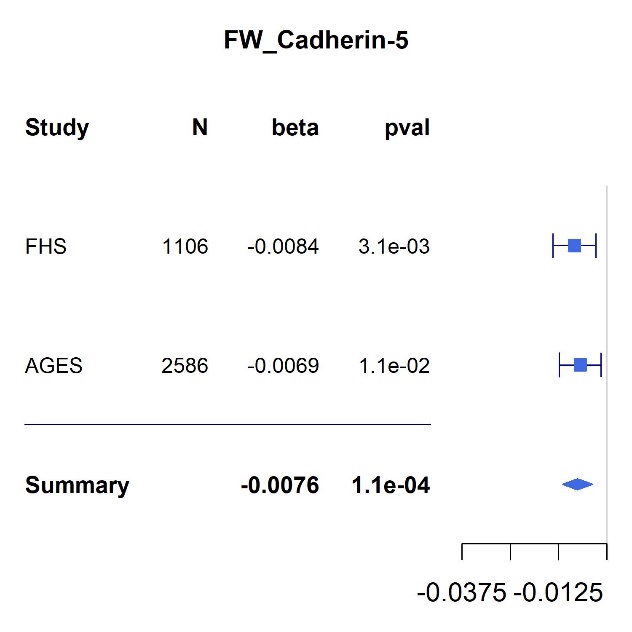

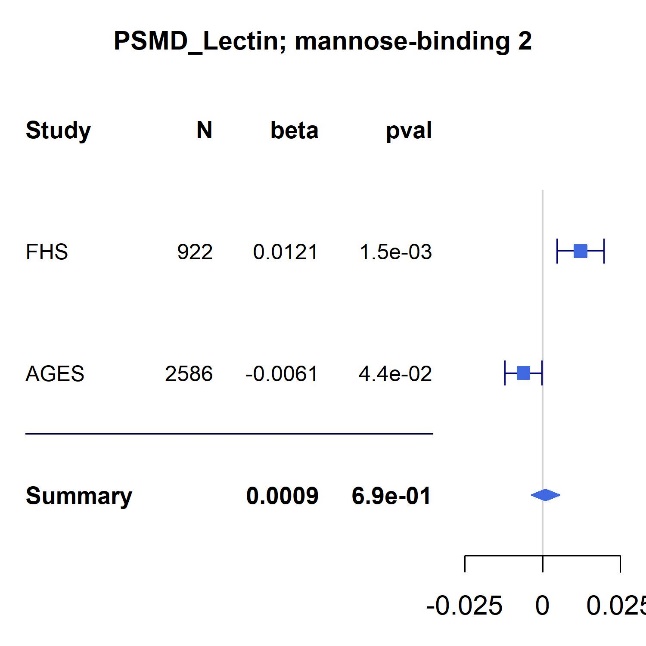

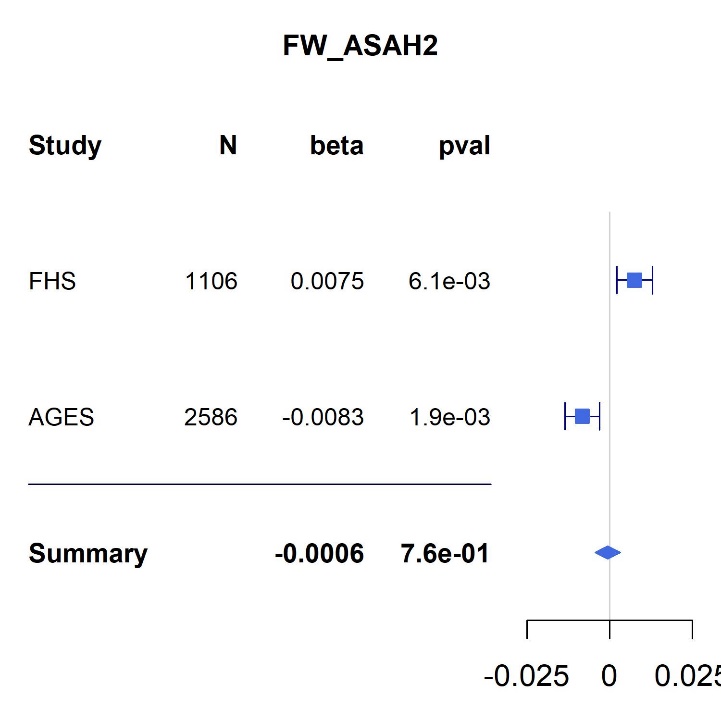

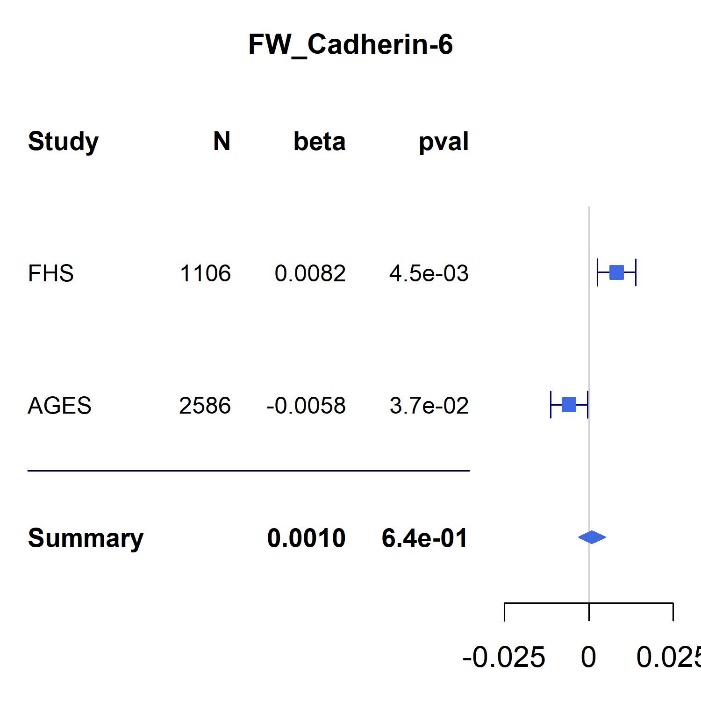

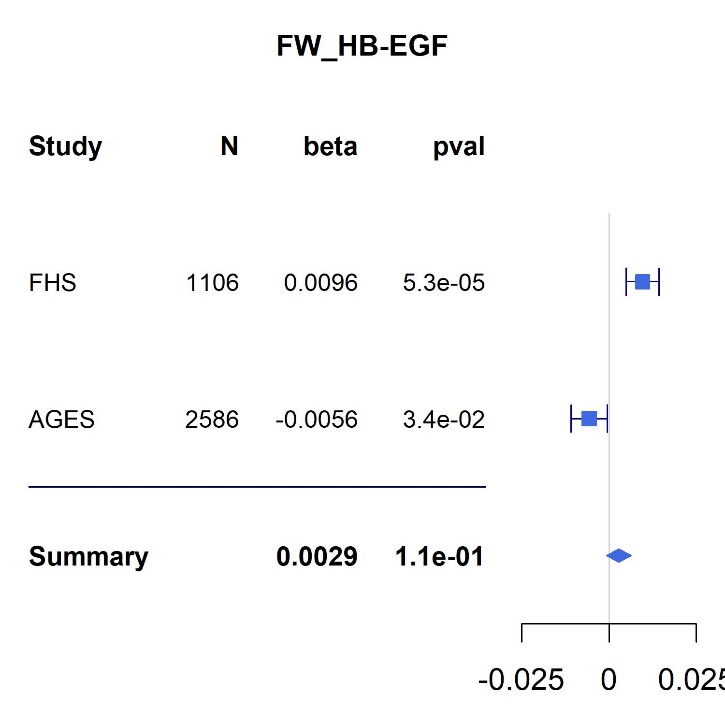

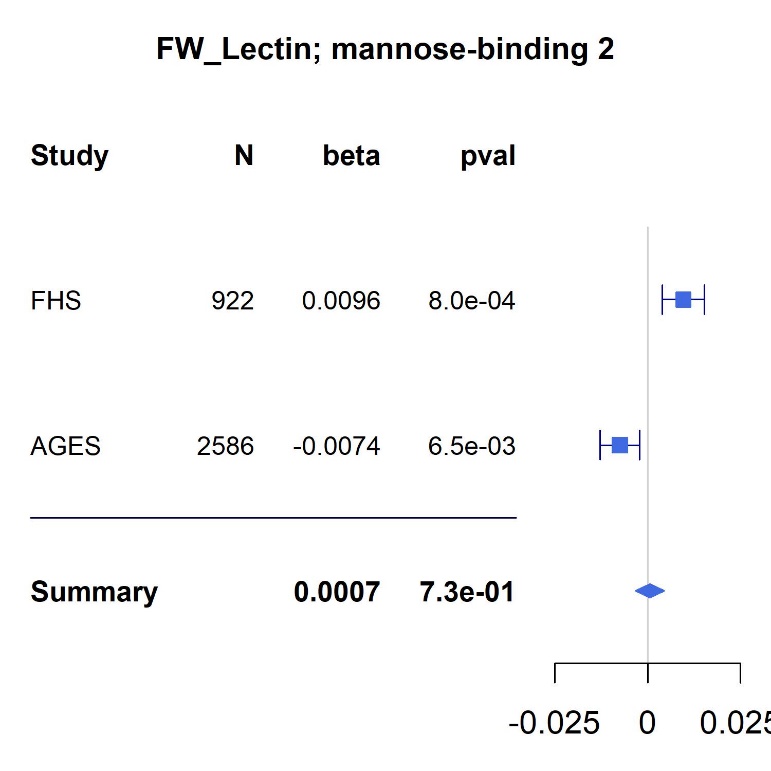

**Figure 2s.** Protein-protein interaction network using the coding genes of top associated proteins with FW (n=23, p<0.01). Each node represents the coding gene of an input protein. Colors represent one of three clusters from k-means clustering analyses. Edges connecting nodes represent known or predicted protein-protein associations.
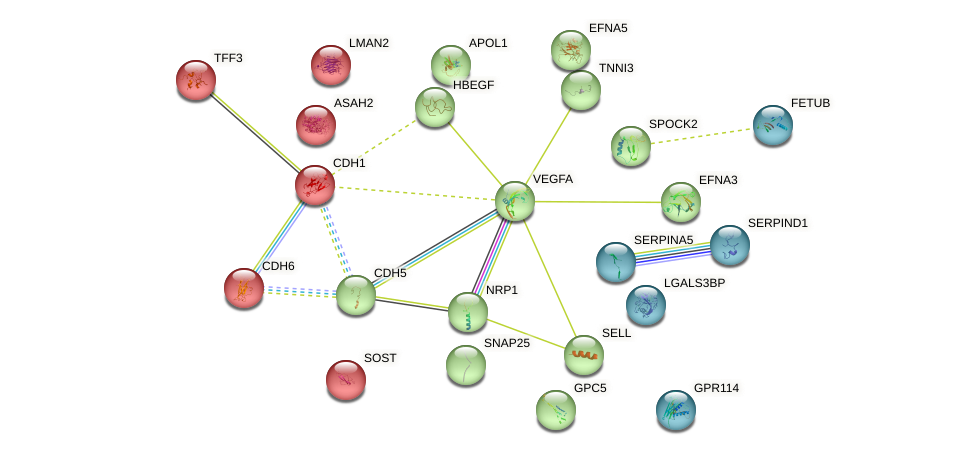

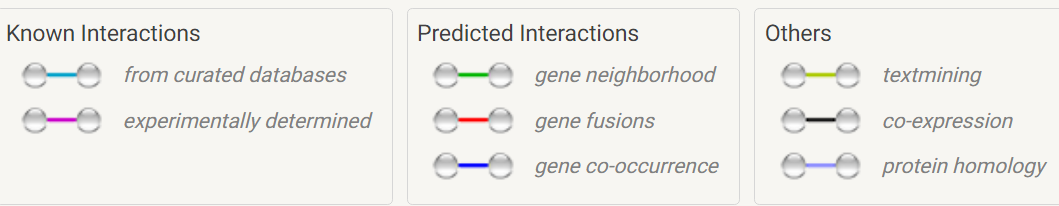

**Figure 3s.** Protein-protein interaction network using the 28 coding genes of the top associated proteins with PSMD (n=25, p<0.01). Each node represents the coding gene of an input protein. Colors represent one of three clusters from k-means clustering analyses. Edges connecting nodes represent known or predicted protein-protein associations.

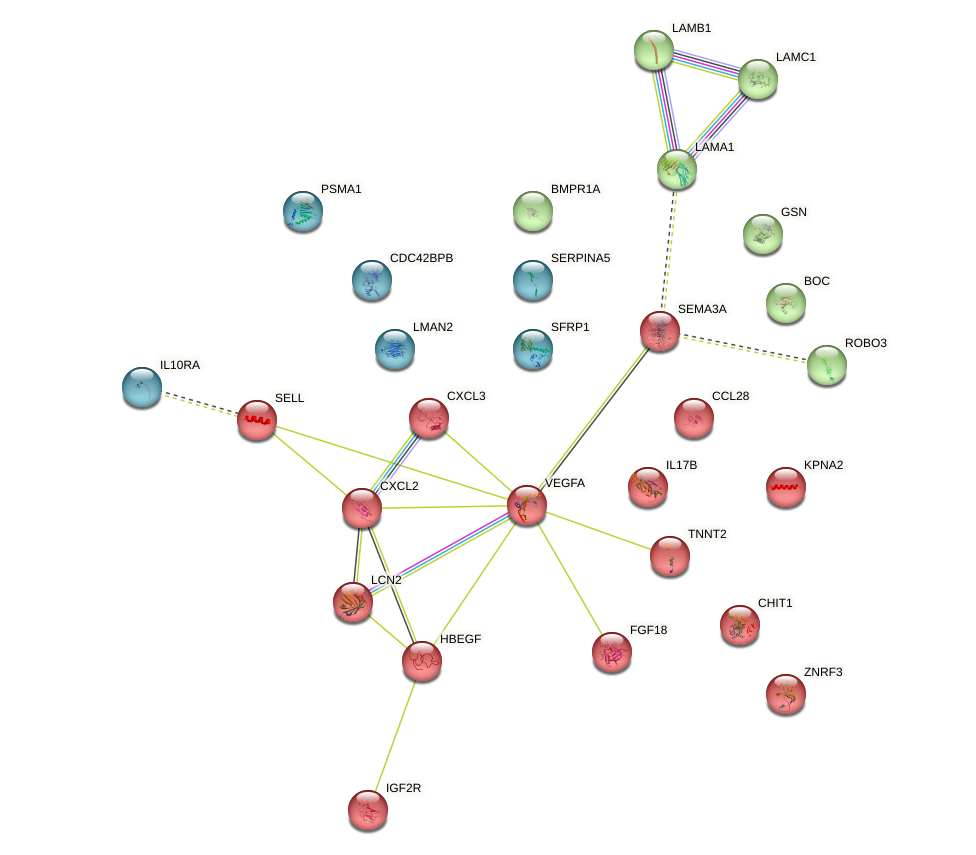

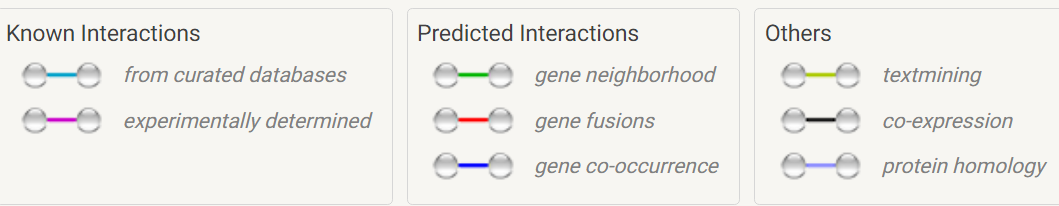

**Figure 4s.** The most significant network generated by IPA using the coding genes of top associated proteins (n=23, p<0.01) with FW.

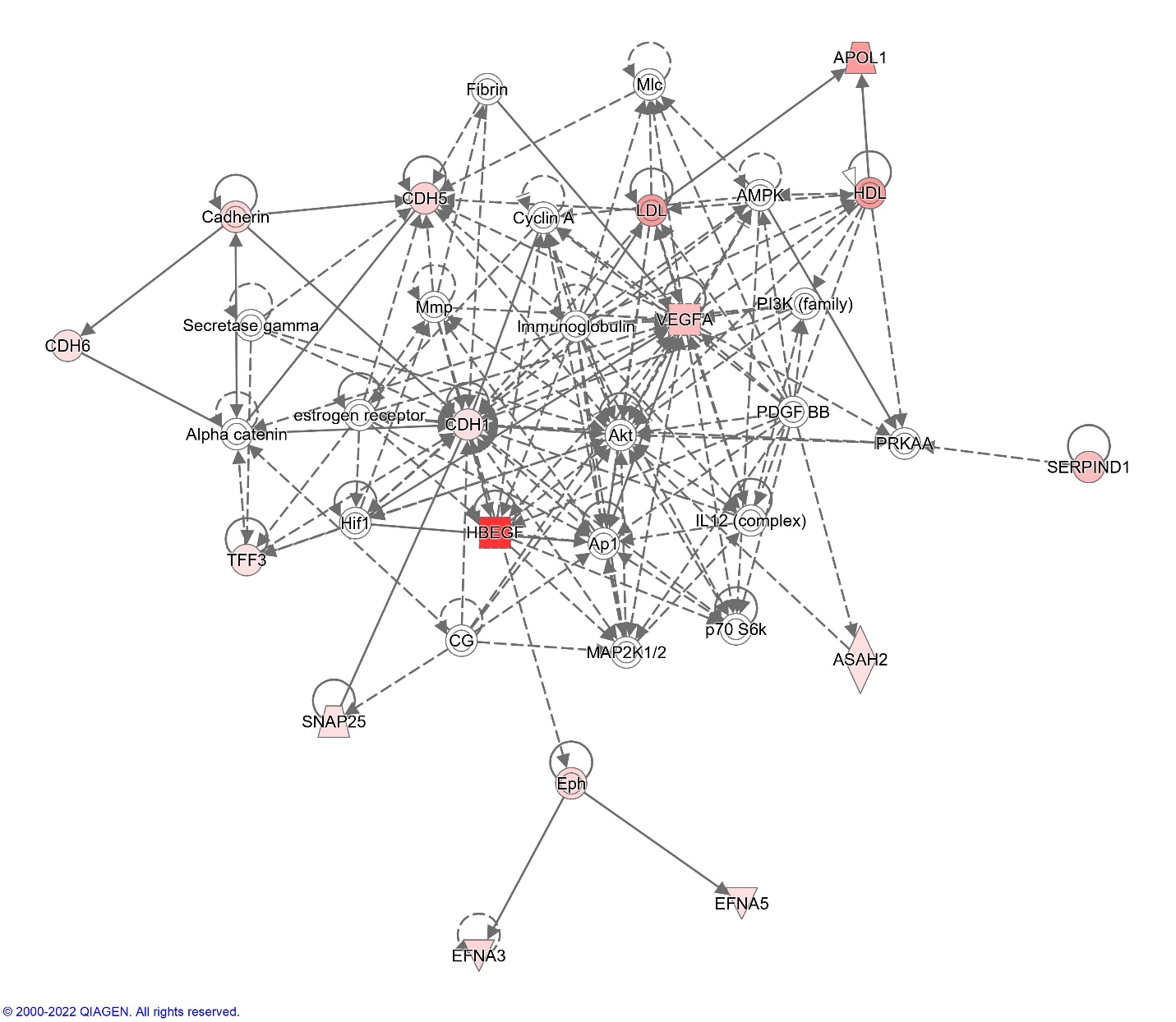

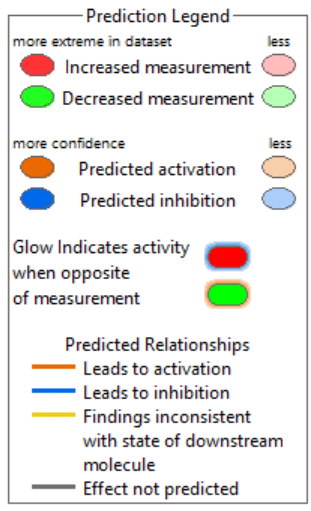

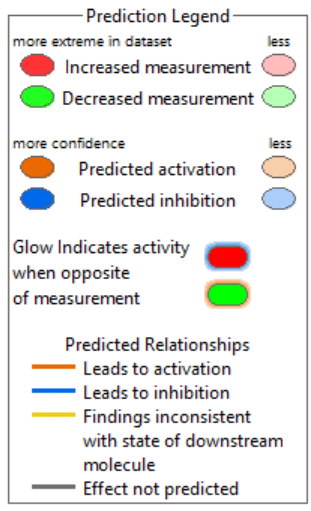

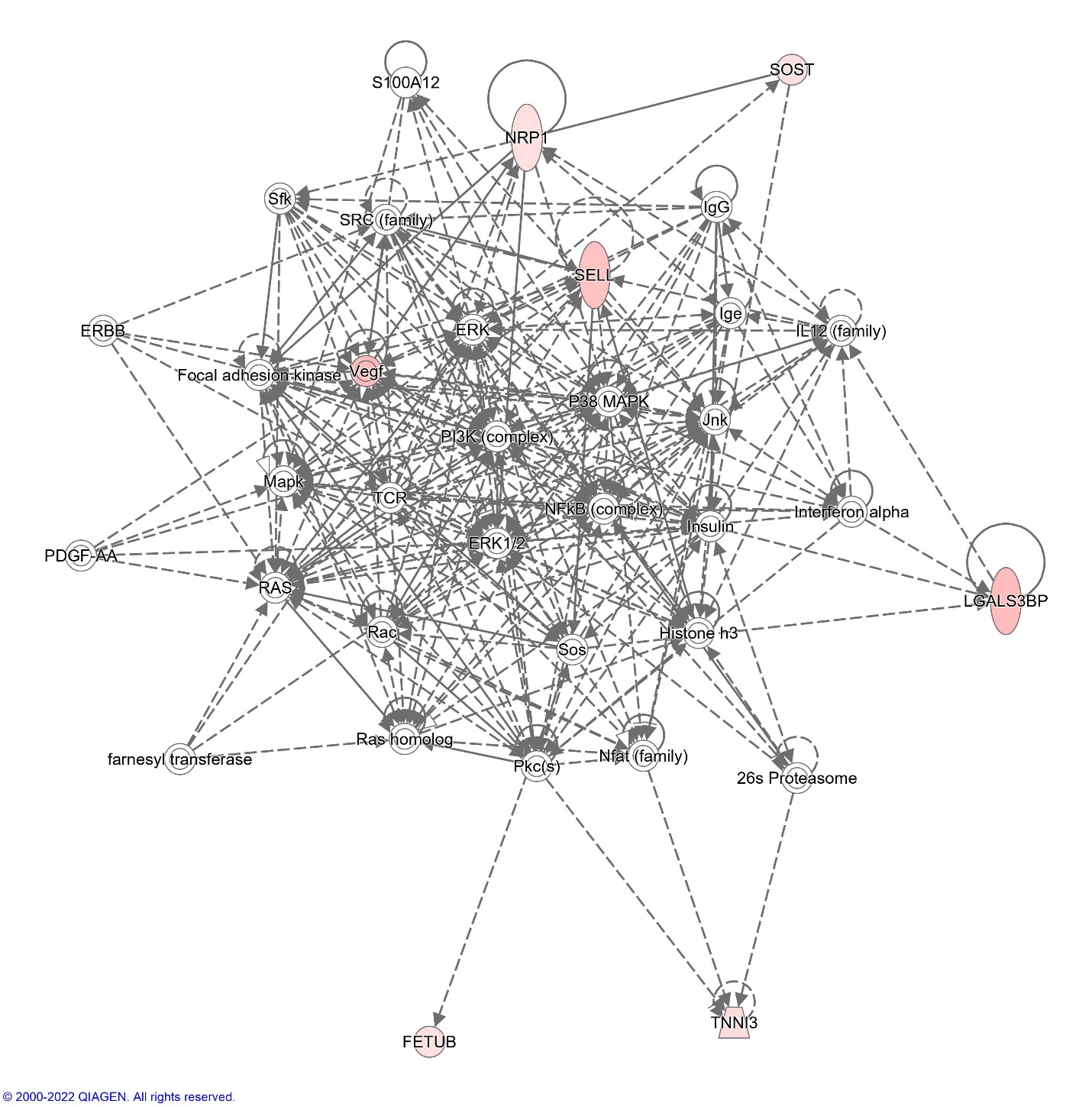
**Figure 5s.** The second most significant network generated by IPA using the coding genes of top associated proteins (n=23, p<0.01) with FW.

**Figure 6s.** The most significant network generated by IPA using the 28 coding genes of the top associated proteins (n=25, p<0.01) with PSMD.

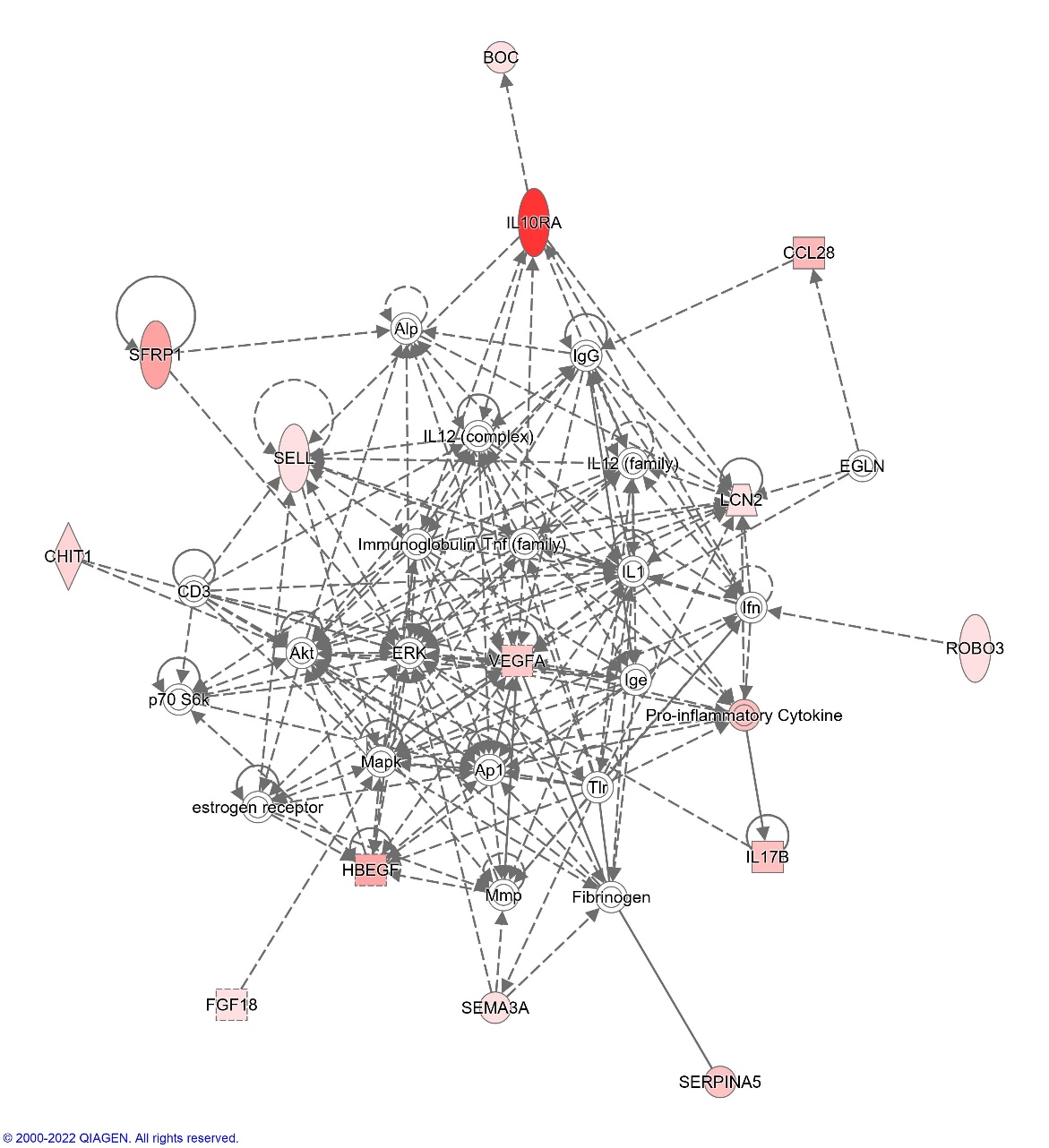

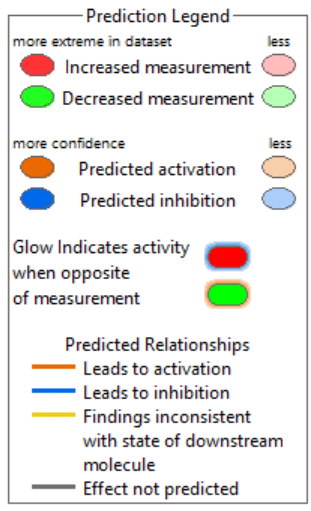

**Figure 7s.** The second most significant network generated by IPA using the 28 coding genes of the top 25 proteins associated with PSMD.

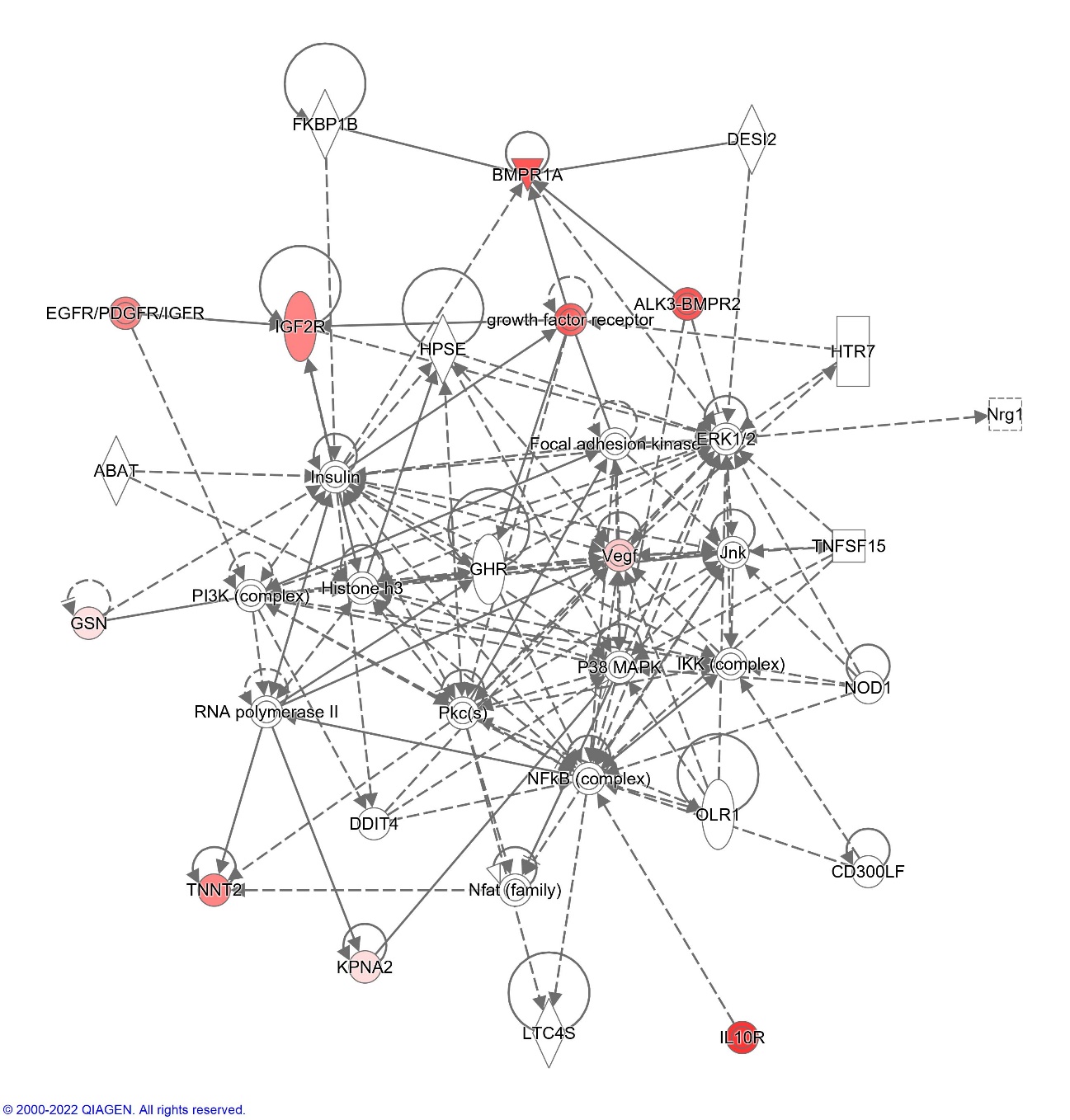

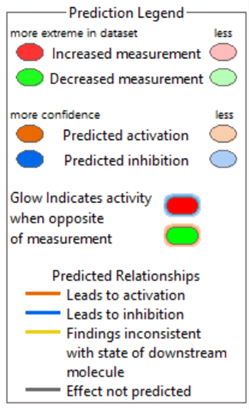
